# Modulated smooth muscle cells accumulate late in human coronary atherosclerosis and are temporally and spatially linked to necrotic core formation

**DOI:** 10.1101/2025.02.18.25322466

**Authors:** Daniel Morales-Cano, Diana Sharysh, Julián Albarrán-Juárez, Antonio de Molina, Verónica Labrador-Cantero, Cecilie Markvard Møller, Laura Carramolino, Jacob F. Bentzon

**Affiliations:** Centro Nacional de Investigaciones Cardiovasculares (CNIC), Madrid, Spain; Department of Clinical Medicine, Aarhus University, Aarhus, Denmark; Department of Cardiothoracic and Vascular Surgery, Aarhus University Hospital, Aarhus, Denmark; Steno Diabetes Center Aarhus and Department of Cardiology, Aarhus University Hospital, Aarhus, Denmark

## Abstract

**Background and Aims:** Proliferation of arterial smooth muscle cells (SMCs) and their modulation to alternative mesenchymal phenotypes is a central mechanism in the growth of atherosclerotic lesions. The underlying processes have been studied extensively in mouse models, but a detailed analysis of when and where modulated SMCs accumulate in human atherosclerosis is lacking. The present study mapped modulated SMC subtypes during the progression of human coronary atherosclerosis and explored their associations with disease processes in human carotid plaques.

**Methods:** Multiplex immunostaining protocols based on single-cell RNA sequencing-validated markers were established to detect SMCs, modulated SMCs, and macrophages in sections of left anterior descending arteries from forensic autopsies. The material comprised 44 arterial segments from 38 individuals, encompassing samples with normal intima, eccentric intimal thickening, pathological intimal thickening, and fibroatheroma. A similar analysis of carotid endarterectomy samples allowed examination of the involvement of modulated SMCs in fibrosis, calcification, and apoptosis. Coronary and carotid sections were analyzed by machine learning-assisted cell classification, enabling phenotyping of entire plaques at high microscopic resolution.

**Results:** Cells co-expressing contractile and modulated SMC markers were present in normal human coronary arteries, but fully modulated SMCs, with complete loss of detectable contractile protein expression, did not accumulate substantially until the fibroatheroma stage, where they were located preferentially around the necrotic core. SMC subtypes showed no preferential co-localization with areas of fibrosis or calcification; however, osteoprotegerin secreted by modulated SMCs was found bound to calcium deposits. Modulated SMCs accounted for 35-53% of all apoptotic remnants for which a cell origin could be determined.

**Conclusions:** Fully modulated SMCs expand at the fibroatheroma stage, localize around the necrotic core region, and account for many apoptotic remnants in plaques.

## Introduction

Smooth muscle cells (SMCs) reside in the walls of many tubular and hollow organs, including arteries, bronchi, intestines, and the bladder, and bestow upon these organs the ability to contract and dilate. Contractile function relies on contractile proteins, which also serve as markers of SMC identity. However, in diseases such as atherosclerosis, SMCs modulate to mesenchymal cells with characteristics of fibroblasts and chondrocytes.^1–3^ These modulated SMCs, often referred to as fibromyocytes, chondromyocytes, or fibrochondrocytes,^1,4–6^ have been the subject of extensive lineage tracing studies in mouse models, and single-cell RNA-sequencing (scRNA-seq) studies have confirmed their presence in human atherosclerosis.^7^ The transcriptional programs of modulated SMCs suggest an involvement in fibrosis and calcification, but little is known about their function. Moreover, it remains unknown at what stage of human atherosclerosis the different modulated SMC subtypes appear, where in the plaque they are located, or what spatial association they show with specific disease processes.

Such an analysis is not trivial. The first requirement is access to human plaque material that encompasses the full pathogenesis of atherosclerosis, including early lesion stages. These samples then need to be examined by multiplex immunostaining for several markers to distinguish modulated SMCs from contractile SMCs and macrophages. Finally, determining the abundance and geographical localization of modulated SMCs in a plaque requires the phenotyping of thousands of cells at high microscopic resolution.

In the present study, we developed multiplex immunofluorescent staining protocols based on alpha-smooth muscle actin (ACTA2), the extracellular matrix proteoglycan lumican (LUM), and the secreted glycoprotein osteoprotegerin (TNFRSF11B) to identify subtypes of SMC-derived cells in autopsy samples of human coronary arteries, covering the range from normal intima to advanced fibroatheromas. We also implemented and validated an automated cell classification technique for accurate (low error rate) phenotyping of all cells in large human plaques. This analysis revealed a graded phenotypic modulation and showed that fully modulated SMCs, with complete loss of ACTA2, expand at the fibroatheroma stage, preferentially localizing around the necrotic core. Analysis of freshly obtained plaques from carotid endarterectomies revealed that fully modulated SMCs account for a high proportion of apoptotic cell remnants.

## Methods

### Human plaque material

Sections (5 µm) of proximal left anterior descending (LAD) coronary artery were obtained from a repository of arterial segments collected during forensic autopsies at the Institute of Forensic Medicine, University of Aarhus, Denmark, between 1996 and 1999.^8^ The samples were obtained from individuals aged 20-80 years, irrespective of the cause of death, with authorization from the Regional Research Ethics Committee and the Danish Data Protection Agency, which complies with the Declaration of Helsinki. The sample collection is anonymized and is not linked to any clinical information except for sex and age group (<45 or ≥45 years). Samples from individuals ≥45 years old were decalcified in 10% formic acid for 24 hours before paraffin embedding. Blocks were selected according to the lesion classification of Dalager et al.^8^ Atherosclerosis stage was confirmed by staining sections with hematoxylin-eosin, with reclassification according to the Virmani classification if needed.^9^ Arteries lacking evidence of disease (foam cells, lipid pools, necrosis, or calcifications) were classified as having normal intima or eccentric intimal thickening (EIT), depending on whether the intima was thinner or thicker than the underlying media. Lesions with lipid pools but no mature necrotic cores were classified as pathological intimal thickening (PIT), and those with a necrotic core as fibroatheromas. This study examined a total of 44 paraffin blocks from 38 individuals, with tissue classified by arterial histology as normal intima (n=11; 6 male and 5 female), EIT (n=11; 6 male and 5 female), PIT (n=12; 8 male and 4 female), and fibroatheroma (n=10; 7 male and 3 female).

For the analysis of associated disease processes (plaque fibrosis, calcification, and apoptosis), fresh carotid plaques (n=12 blocks from 8 females) were collected from endarterectomies conducted at the Department of Vascular Surgery, Aarhus University Hospital. These samples were fixed in formaldehyde for 24 hours immediately upon collection and then embedded in paraffin, and 5 µm sections were cut for the present study. As the samples were anonymized and collected during standard surgeries (not for this study), ethics board approval was not required (Danish Act no. 593 of 14 June 2011 on Research Ethics Review of Health Research Projects).

### Single-cell RNA-seq dataset analysis to identify modulated SMC markers

To define modulated SMC-derived cell populations and their markers, we analyzed the publicly deposited single-cell RNA sequencing data for human coronary plaques from Wirka et al. and for carotid endarterectomies from Pan et al. and Alsaigh et al.^1,2,6^ The datasets (GSE131778, GSE155512, and GSE159677) were downloaded as FASTQs and processed with CellRanger (v6.1.1) and the hg38 human reference genome (Ensembl 109) to reduce preprocessing differences. Data were then analyzed in Seurat version 4.0.4 running in R version 4.2.2.^10^ The parameters for the initial filtering were as follows: detected genes, < 4000; 300 < UMIs < 30000; and <10% mitochondrial, <60% ribosomal, and <1% hemoglobin genes expressed. Doublets were removed with DoubletFinder, assuming a 15% doublet rate.^11^ Datasets were normalized individually with SCTransform and integrated using the CCA Seurat method.^12^ Coronary and carotid plaque cells were analyzed separately after additional data cleaning to retain only cells with the following features: detected genes, 1100–4000 (coronary), 900–4000 (carotids); 1300 < UMIs < 20000. Principal component analysis included 40 principal components, and UMAP embedding was run with 22 (coronaries) or 18 (carotids) principal components, followed by clustering. The clusters were manually annotated based on the expression of known marker genes (PTPRC, TYROBP, PECAM1, PI16, and ACTA2), and mesenchymal cell types were selected and reclustered. Cells expressing *PTPRC* (encoding CD45) or *TYROBP* were defined as contaminants from other clusters and removed. A pseudotime trajectory was inferred for coronary plaque data using Slingshot (dynverse suite) with the root in the contractile SMC cluster.^13^ The expression levels of selected genes (*ACTA2, TNFRSF11B*, and *LUM*) were then analyzed along the pseudotime trajectory branch towards fibroblasts. To determine the significance of inferred pseudotemporal changes in the expression of these genes, we fitted a generalized additive model (using the ‘GAM’ R package) with pseudotime as an independent variable and locally estimated scatterplot smoothing (LOESS) of gene expression as a dependent variable. To test the similarity of mesenchymal-cluster diversity and cell type composition in the coronary and carotid datasets, we transferred cluster names for coronary SMCs and modulated SMCs to the carotid dataset and projected the carotid plaque data onto the UMAP structure of the coronary plaque data using the Seurat functions TransferData and MapQuery, respectively.^12^

### Multiplex immunostaining

Slides were dewaxed, rehydrated, and transferred to 0.05 % Tween 20 citrate buffer (10 mM, pH6.0). To retrieve antigens, slides were then heated for 3 min in a pressure cooker (LAD sections) or a microwave oven at 350W for 5 min (endarterectomy sections). After two washes with phosphate-buffered saline (PBS), sections were permeabilized by incubation for 10 min at room temperature in PBS, 0.3% Triton X-100 and then blocked for 1 h at room temperature with 10% normal goat serum (Life Technologies, 16210-072) in PBS, 0.1% Tween20. Blocked sections were incubated overnight at 4°C with combinations of primary antibodies (details in **Supplementary Table 1**), followed by washes and incubation for 1 h at room temperature with matched fluorophore-conjugated secondary antibodies (details in **Supplementary Table 2**). Nuclei were stained with DAPI (Sigma-Aldrich 1.24653), and slides were mounted in SlowFade Antifade mounting medium (Invitrogen S36937). Adjacent sections were incubated with isotype control antibodies at the same concentrations used for the corresponding primary antibody incubations.

### Analysis of established cell type markers in LAD sections

Confocal images of CD45/CD68/ACTA2-stained fibroatheroma sections were acquired with a Nikon AR1 confocal microscope at 40x magnification using a Plan Fluor 40x/1,3 Oil DIC H N2 Oil objective. Z-stack images (z-step 1 μm) were acquired from each lesion in predefined regions, including plaque shoulders, cap (sides and center), and core border (sides and center). Each image was collapsed into a maximal intensity projection, and DAPI-stained nuclei were identified with a macro in Fiji (ImageJ distribution).^14^ Each nucleated cell profile was then manually characterized for expression of CD45, CD68, and ACTA2.

### Analysis of modulated cell-type markers in whole plaque sections

LAD and carotid endarterectomy sections were stained with CD68/ACTA2/LUM or CD68/ACTA2/TNFRSF11B antibody combinations and scanned with an AxioScan Z1 slide scanner at 20x magnification using a Plan-Apochromat 20x/0.8 M27 objective. Fiji (ImageJ distribution) was used to adjust brightness and contrast and to remove autofluorescence with the AFID plugin developed by Baharlou et al.^14,15^ Identical microscope settings and image processing parameters were used to image adjacent isotype control-stained sections in the same session.

Counting of cell phenotypes (marker expression profiles) across whole plaque sections was automated by training a cell classifier tool in QuPath (v0.2.3 and 0.3.2).^16^ Briefly, the Random Trees machine-learning algorithm was trained on 300-500 human-classified cells in 10 shoulder regions in 5 coronary fibroatheromas (for coronary lesion analysis) and 20 randomly selected regions in 10 endarterectomy sections (for carotid lesion analysis). The trained classifiers for each marker were then applied sequentially to phenotype all plaque cells in the coronary or endarterectomy samples. The machine learning analysis was validated by comparing its results with manual cell counts by linear regression for 5 complete coronary fibroatheromas (containing approximately 12000 nucleated cell profiles in total).

Cell types were counted throughout the intima or plaque in all sections. In LAD fibroatheromas, cell types were also quantified in specific plaque regions of interest (ROIs), defined according to a previously described plaque subdivision.^17^ Operators manually delineated the lumen, plaque (excluding underlying media), and the necrotic core (the plaque region devoid of cells except for scattered nuclei) and selected the two points on the luminal surface at the junction between non-plaque and plaque tissue. These coordinates set the limits for automated delineation of the *border zone* (the 100-µm-deep plaque region surrounding the necrotic core), *shoulder regions* (plaque tissue ≤750 µm from the luminal points at the plaque–non-plaque junction), and the *luminal region* (non-shoulder plaque tissue at a depth of ≤200 µm from the lumen surface). To ensure that plaque tissue was assigned to only one ROI, areas fulfilling more than one criterion were assigned according to the following order of precedence: border zone, shoulder, luminal region. Plaque tissue not included in any of these ROIs was defined as *other*. In this way, the sum of the ROIs and the necrotic core area equaled the total plaque area.

### Analysis of apoptosis in endarterectomy samples

To identify and phenotype apoptotic cell remnants, sections of endarterectomy samples were stained using TUNEL (terminal deoxynucleotidyl transferase-mediated dUTP nick end labeling) with the In Situ Cell Death Detection Kit, Fluorescein (11684795910, Sigma) in combination with CD68/ACTA2, CD68/LUM, or CD68/TNFRSF11B immunofluorescence. Sections were counterstained with DAPI and scanned in an epifluorescence Leica DM6B upright microscope using the tile-scan Navigator module with an HCL APO 10x/0.3 W dipping objective. To confirm specificity, all acquisition and postprocessing settings were tested on sections stained with isotype control antibodies. Fiji was used to adjust brightness and contrast and to reduce autofluorescence from marker channels by subtracting an autofluorescence image obtained in an empty channel. Labeled cells were then automatically counted using QuPath version 0.2.3, as described above.

### Alizarin Red and Aniline Blue staining

Endarterectomy sections were stained using standard protocols with Alizarin Red or Aniline Blue (as a component of Masson’s trichrome) to detect calcified and fibrous tissue, respectively. Masson’s trichrome staining was performed on ACTA2/LUM/CD68-stained sections after AxioScan Z1 acquisition and removal of the coverslip. Because antigen retrieval in citrate buffer (pH 6) removes calcium deposits, Alizarin Red staining was performed on adjacent sections to those stained for ACTA2/CD68/TNFRSF11B. Brightfield images were digitalized with an AxioScan Z1 slide scanner at 20x magnification using a Plan-Apochromat 20x/0.8 M27 objective. ROIs covering Aniline Blue- and Alizarin Red-stained areas were defined with the color deconvolution and color threshold plugins, respectively in Fiji and transferred to corresponding ACTA2/CD68/LUM and ACTA2/CD68/TNFRSF11B images. The proportions of cell types inside and outside ROIs were then determined by automated cell counting in QuPath version 0.2.3, as described above.

### Statistics

Cell fractions of each staining profile within plaques or plaque ROIs were compared across categories (plaque region, arterial morphology, and associated disease processes) using the Kruskall-Wallis test followed by Dunn’s post hoc test, comparing either all categories or using a specific category as the comparator, as specified in the figure legends. Agreement between automated and manual cell counts was tested by linear regression. Calculations were performed in Prism 9 (GraphPad Software), and differences were considered statistically significant at P < 0.05. Statistical analysis for single-cell RNA-seq data is described above.

### Data and code availability

The code used to analyze public scRNA-seq data, ImageJ macros, and a step-by-step protocol with an associated macro for automated cell-type phenotyping with Qupath version 0.3.2. can be downloaded from https://github.com/LAB-JFB. All scanned images are available upon reasonable request.

## Results

### Abundance and localization of modulated SMCs in coronary fibroatheromas

To map modulated SMCs at different stages of coronary atherosclerosis, we examined a collection of LAD samples obtained from forensic autopsies of individuals aged 20–80 years, encompassing the full natural progression of lesion development.^8^

We first estimated the size and location of the modulated SMC population in advanced coronary atherosclerotic plaques using established cell-type markers. Coronary fibroatheromas (n=10) were examined by multiplex immunostaining to identify contractile SMCs (ACTA2+), macrophages (CD68+), and lymphocytes and other hematopoietic cells (CD45+), with triple-negative cells being considered putative modulated SMCs (**Fig. 1A-C**). In each section, we analyzed 8 regions per plaque at high magnification, including the two plaque shoulders, three areas in the luminal region, and three areas in the necrotic border region. ACTA2+ SMCs were mostly located in the luminal and shoulder regions, whereas triple-negative cells (putative modulated SMCs) constituted almost half of the cells near the necrotic core (**Fig. 1D**). We also detected a large fraction of CD68+ cells not expressing CD45+. These could be CD68+ modulated SMCs or macrophages with no detectable CD45 expression.

**Figure 1.**
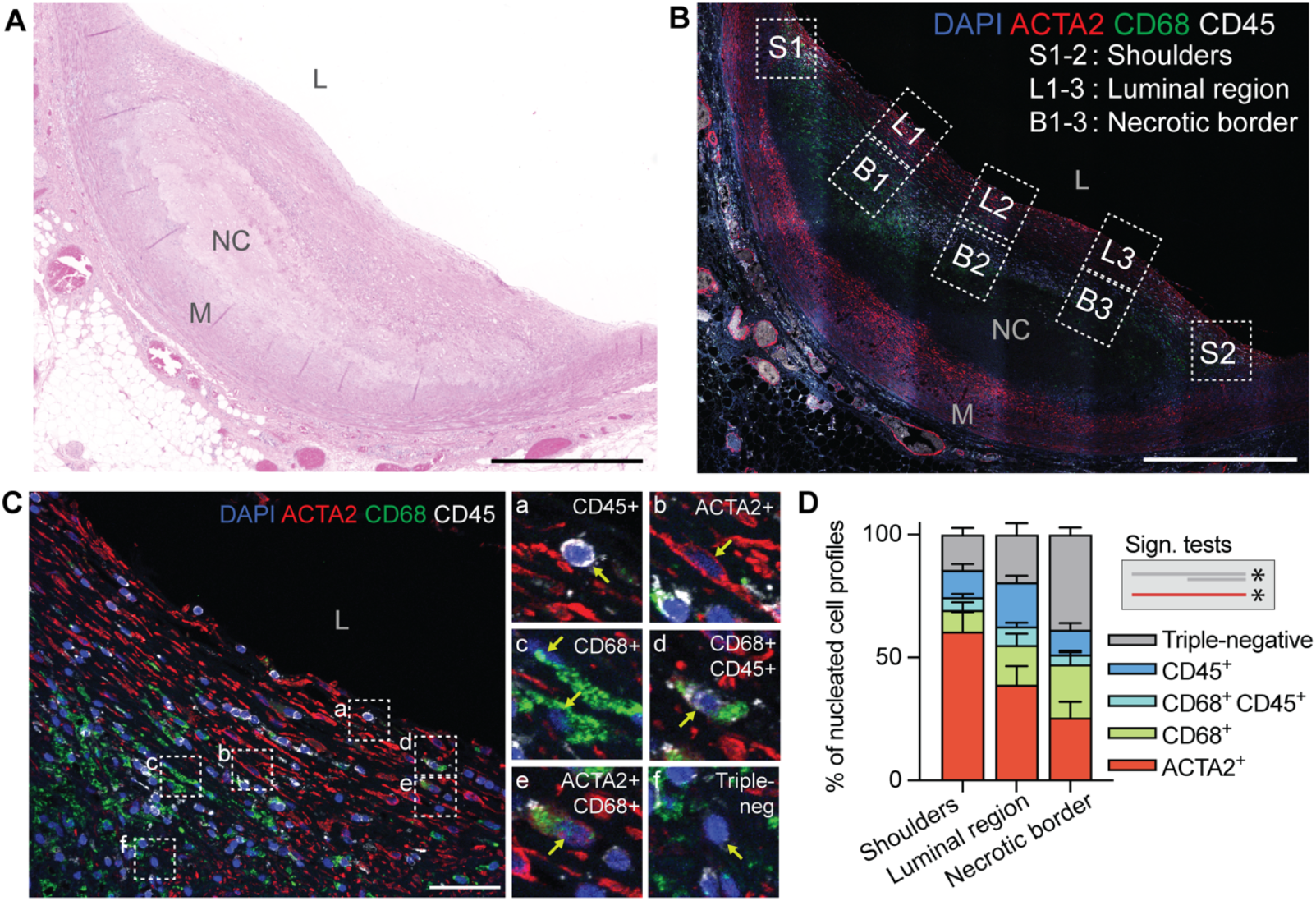
Rule-out approach with established cell markers to identify modulated SMCs in coronary fibroatheromas. **A,** Representative hematoxylin–eosin-stained fibroatheroma from the human autopsy LAD plaque library. Scale bar, 1 mm. **B,** The same plaque immunostained for ACTA2, CD68, and CD45. Scale bar, 1 mm. Boxes demark regions analyzed at high magnification in C. **C,** Shoulder region (S2) from the same plaque, showing examples of CD45+ hematopoietic cells (a), ACTA2+ SMCs (b), CD68+ cells (c), CD45+CD68+ macrophages (d), rare ACTA2+CD68+ cells (e), and cells negative for all three markers (f), identifying them as putative modulated SMCs. Scale bar, 50 μm. **D,** Cellular composition in different plaque regions (n=10 fibroatheromas analyzed). ACTA2+ cells predominate in the shoulder and luminal regions, whereas putative modulated SMCs (ACTA2/CD45/CD68-negative) are abundant in the necrotic core border. Bars indicate mean ± SEM. *P<0.05 (comparisons indicated by line color and length) by Kruskall-Wallis and Dunn’s post hoc test. L, Lumen; M, Media; NC, necrotic core.

### Markers of modulated SMCs identified from scRNA-seq data

Previous scRNA-seq studies of advanced human coronary atherosclerosis reported lumican (*LUM*) and osteoprotegerin (*TNFRSF11B*) as candidate marker genes of modulated SMCs with fibroblast-like and osteochondrogenic-like phenotypes, respectively.^1,4^ To assess the performance of these markers, we first re-analyzed the coronary plaque scRNA-seq data presented by Wirka et al.^1^ The mesenchymal cell supercluster, comprising pericytes, contractile SMCs, modulated SMCs, and fibroblasts, was isolated and re-clustered, and the expression of *ACTA2, LUM*, and *TNFRSF11B* was analyzed across the main axis of phenotypic diversity running from contractile, *ACTA2-*expressing SMCs to fibroblasts (**Fig. 2A-B**). *TNFRSF11B* was expressed mainly by cells in the transition zone between *ACTA2*-expression and *ACTA2*-nonexpression, whereas *LUM* was expressed by fully modulated SMCs lacking *ACTA2* expression. To formally characterize these patterns, we used Slingshot to calculate a pseudotime trajectory running from contractile SMCs to fibroblasts, and then correlated pseudotime coordinates to the expression of *ACTA2, TNFRSF11B*, and *LUM* (**Fig. 2C-D**). This analysis verified that *ACTA2, TNFRSF11B*, and *LUM* label sequential cell populations with phenotypes along the main axis of diversity from contractile SMCs toward fibroblasts. Together, these markers label most SMCs and modulated SMCs in plaques and are little expressed in non-mesenchymal cells (**Suppl. Fig. 1**), making them good markers for mapping the accumulation of modulated SMCs in human atherogenesis.

**Figure 2.**
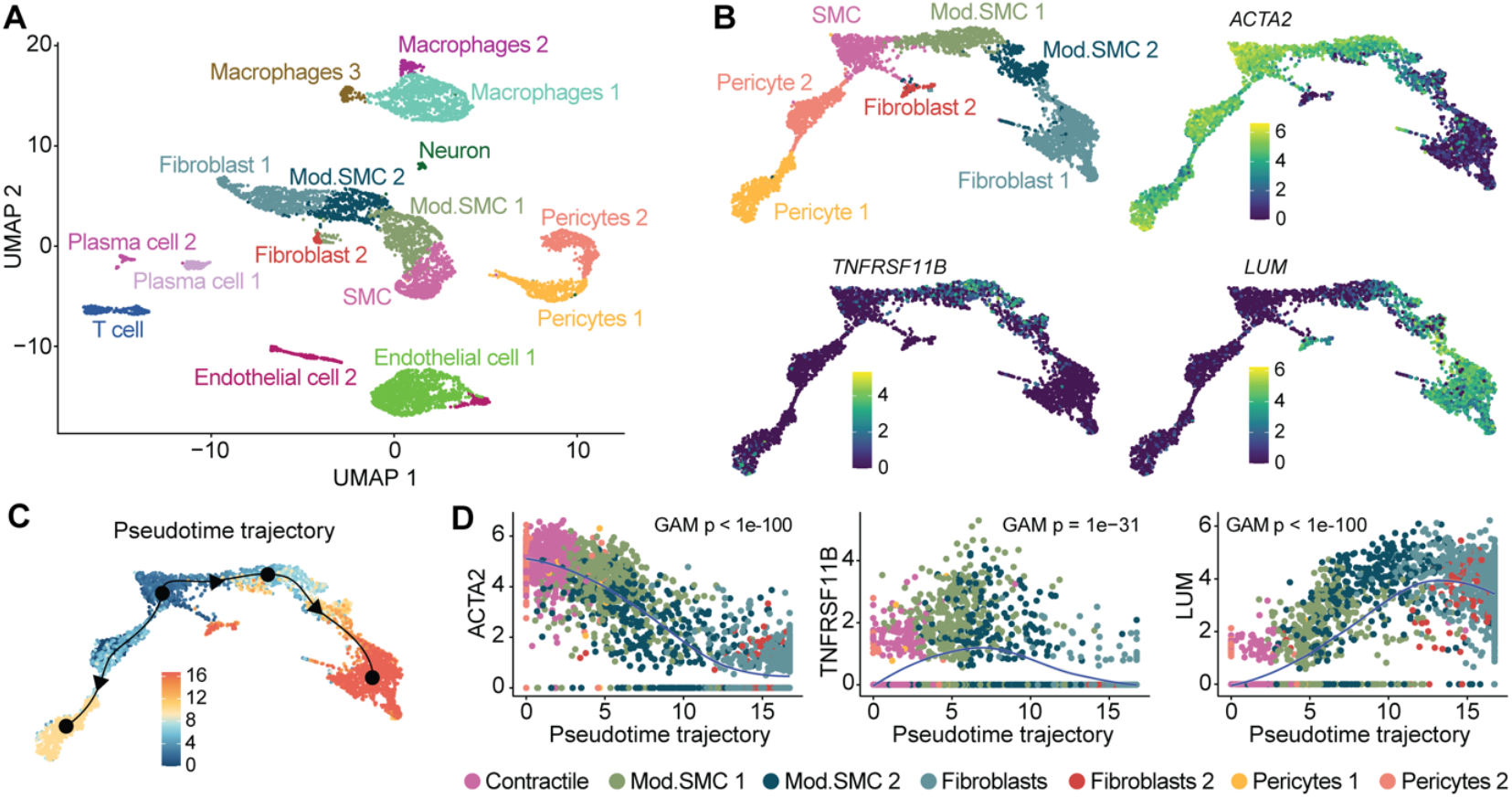
Marker analysis of publicly deposited single-cell RNA seq data from coronary artery plaques. **A,** Clustering and cell-type annotation of the scRNA-seq data from human atherosclerotic coronary arteries published by Wirka et al.^1^ **B,** Visualization of marker gene expression in reclustered mesenchymal cell types (pericytes, SMCs, modulated SMCs, and fibroblasts). Color scale is normalized gene expression. **C,** Inferred pseudotime trajectories from SMCs calculated with the Slingshot tool. Black dots denote ‘milestones’; arrows indicate the trajectory direction towards fibroblasts or pericytes. Color scale is pseudotime coordinates. **D,** Normalized expression of selected genes (ACTA2, TNFRSF11B, and LUM) ordered by inferred pseudotime trajectory (SMC-to-fibroblast branch) and fitted with LOESS regression (blue curve). The association of gene expression with pseudotime coordinates was highly significant in a generalized additive model (GAM).

### TNFRSF11B+ and LUM+ cells expand at the fibroatheroma stage of plaque development

To determine if cells expressing TNFRSF11B or LUM are present in normal intima or accumulate during atherosclerotic plaque formation, we established multiplex staining protocols for ACTA2/CD68/TNFRSF11B and ACTA2/CD68/LUM antibody combinations and stained LAD sections exemplifying normal intima (NI, n=12), eccentric intimal thickening (EIT, n=11), pathological intimal thickening (PIT, n=12), and fibroatheromas (FA, n=10) (**Fig. 3A-B**; isotype control staining in **Suppl. Fig. 2)**. Despite both TNFRSF11B and LUM being secreted proteins, we found clear cellular staining of both markers. Indeed, the strongest signal with the anti-LUM antibody was within cells, suggesting that it binds better to the core protein, before GAG-chain modification. To phenotype all plaque cells at single-cell resolution (a requirement for determining the cellular co-localization of markers), we then trained machine-learning-based tools in Qupath to automatically classify cell phenotype across the entire healthy intima or plaque. The tool was validated by comparison with approximately 12000 manually curated cell types in fibroatheromas stained for ACTA2/CD68/TNFRSF11B or ACTA2/CD68/LUM (n=5 each). Linear regression between machine-learning and manual determinations of cell fractions showed a high level of agreement for all analyzed cell types (**Suppl. Fig. 3**).

**Figure 3.**
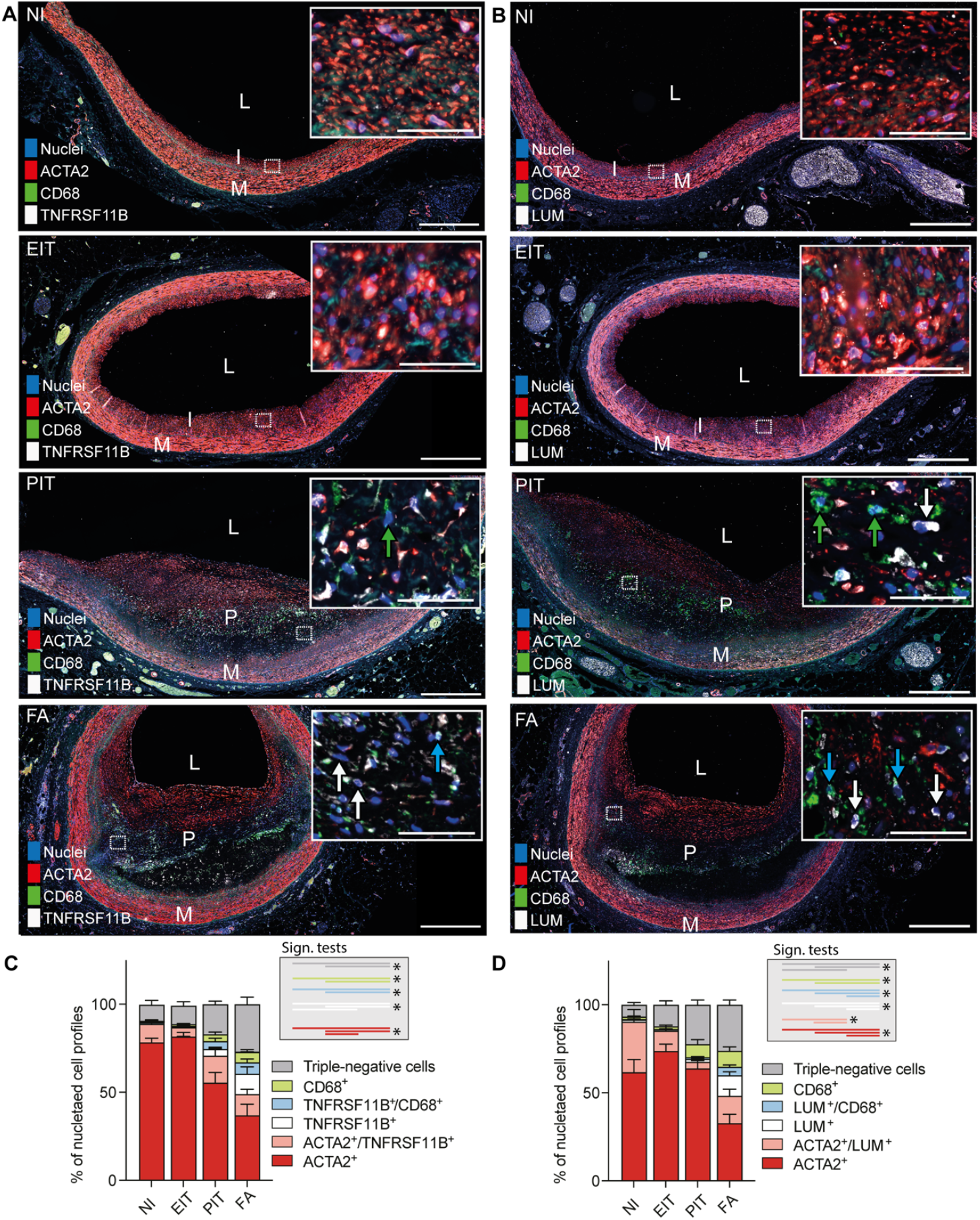
Fully modulated SMCs expand at the fibroatheroma stage. **A-B,** Representative examples of ACTA2/CD68/TNFRSF11B and ACTA2/CD68/LUM staining in coronary artery sections with normal intima (NI, n=11), eccentric intimal thickening (EIT, n=11), pathological intimal thickening (PIT, n=12), and fibroatheroma (FA, n=10). Insets show cells at higher magnification. Arrows in PIT and FA panels point to examples of marker-expressing cells colored according to the code in the graphs in C and D. Scale bars, 500 μm in overviews and 50 μm in insets. **C-D,** Cell phenotype (marker expression profile) according to lesion stage, showing that fully modulated TNFRSF11B+ or LUM+ cells, which have lost expression of ACTA2, accumulate at the fibroatheroma stage. Bars show mean ± SEM. *P<0.05 for the indicated comparisons by Kruskall-Wallis test followed by Dunn’s post hoc test. L, Lumen; M, Media; P, plaque; I, Intima.

The results of automated cell phenotyping are shown in **Fig. 3C-D**. During lesion pathogenesis, there was a decrease in the fraction of ACTA2+ SMCs (with or without concomitant expression of other markers), whereas the numbers of CD68+ macrophages increased. Cells lacking both these markers also increased in abundance, and the identity of these cells could be partly resolved with the LUM and TNFRSF11B markers. Double-positive cells, expressing ACTA2 plus TNFRSF11B or ACTA2 plus LUM, were present in normal intima and throughout lesion development, but modulated cells, expressing TNFRSF11B or LUM but not ACTA2, were restricted to lesions and were substantially more abundant at the fibroatheroma stage, accounting respectively for 12% and 13% of all plaque cells. Interestingly, the ACTA2-negative TNFRSF11B+ and LUM+ populations included cells that also expressed CD68. In the human plaque scRNA-seq data, the specificity of *TNFRSF11B* and *LUM* expression for mesenchymal plaque cells is higher than the specificity of *CD68* expression for macrophages (**Suppl. Fig. 4**), suggesting that TNFRSF11B+CD68+ and LUM+CD68+ cells are more likely to be modulated SMCs than TNFRSF11B+ or LUM+ macrophages, a conclusion supported by the lack of the hematopoietic marker CD45 in many CD68+ cells in fibroatheromas. The appearance of LUM+ cells at the fibroatheroma stage was confirmed for both males and females separately; however, the study was not statistically powered to explore possible sex-specific differences (**Suppl. Fig. 5**).

### LUM+ cells are enriched around the necrotic core

To investigate the spatial distribution of the identified modulated cells in fibroatheromas, we segmented lesions into shoulders, luminal region, necrotic core border, and other regions (**Fig. 4A**). Fully modulated LUM+ cells were significantly more abundant in the necrotic core border than in the luminal region of the plaque (**Fig. 4B-E**). TNFRSF11B+ cells were similarly more abundant near the necrotic border, but in this case the difference did not reach statistical significance. Notably, the spatial association of LUM+ cells with the necrotic core is consistent with our observation that the accumulation of these cells coincides with necrotic core formation, when PIT lesions, lacking a necrotic core, transform into fibroatheromas.

**Figure 4.**
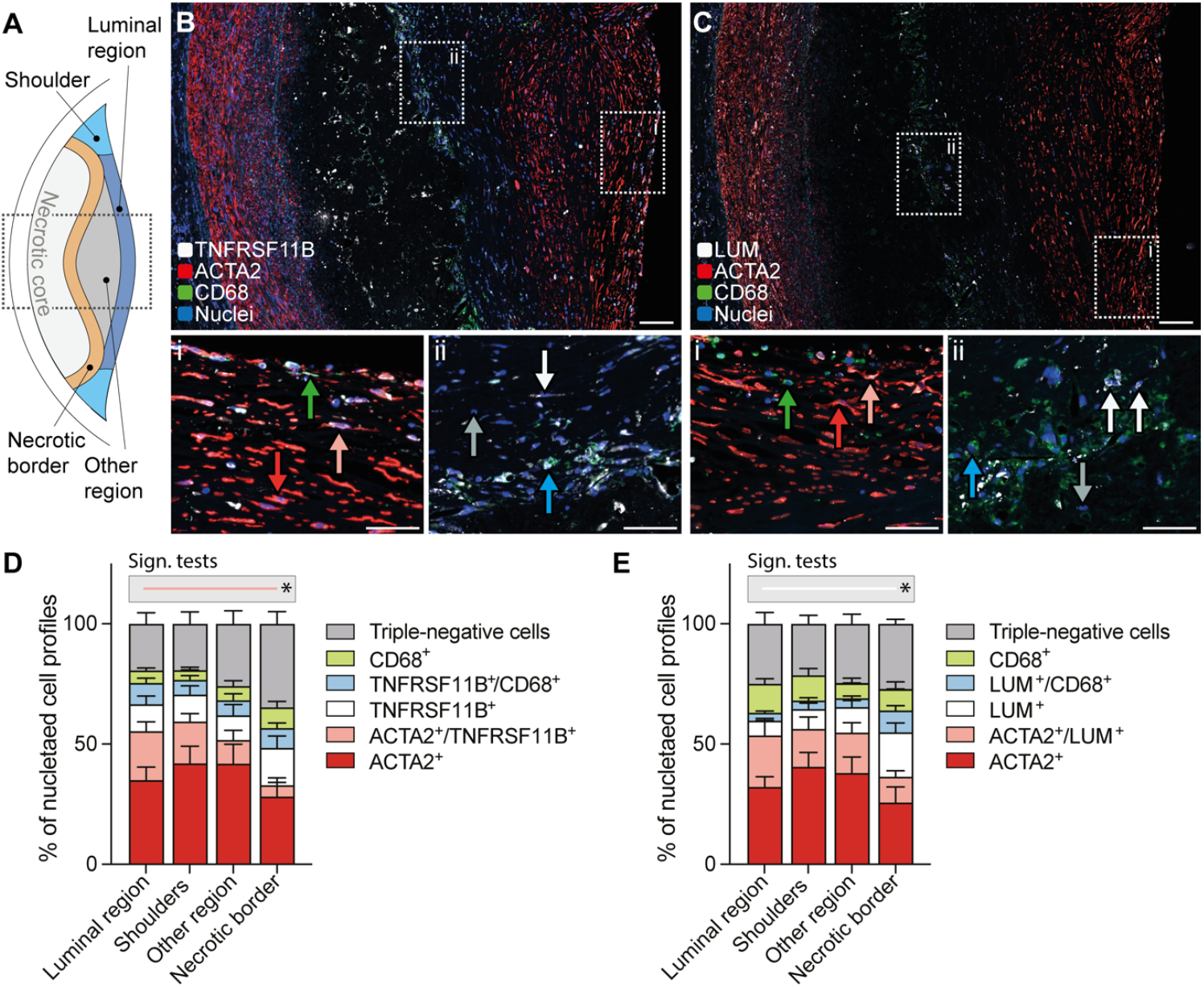
Fully modulated SMCs locate to the necrotic border region. **A,** Schematic showing the fibroatheroma subdivisions in which cellular composition was analyzed (see Methods for details). The boxed area corresponds to the sections shown in B-C. **B-C,** Representative sections immunostained for ACTA2/CD68/TNFRSF11B and ACTA2/CD68/LUM, showing the differences in cellular composition along an axis from the lumen to the necrotic core. Arrows indicate examples of marker-expressing cells colored according to the code in the graphs in D and E. Scale bars, 100 μm in overviews and 50 μm in high magnification images. **D-E,** Cell phenotype across plaque regions, showing that fully modulated LUM+ cells are significantly more abundant in the necrotic border region than in the luminal region (n=10 fibroatheromas analyzed). Bars show mean ± SEM. *P<0.05 for the indicated comparison by Kruskall-Wallis and Dunn’s post hoc tests.

### Association of LUM+ and TNFRSF11B+ modulated SMCs with fibrosis, calcification, and apoptosis

The extracellular-matrix proteoglycan LUM is involved in fibroblast-mediated deposition of fibrillar collagen,^18^ whereas the osteoblast-secreted soluble receptor glycoprotein TNFRSF11B (osteoprotegerin) protects bone from osteoclast resorption.^19^ These actions, coupled with the co-appearance and intraplaque location of cells expressing these markers near the necrotic core, suggested three possible implications in plaque development: an association of LUM+ cells with plaque fibrosis; an association of TNFRSF11B+ cells with plaque calcification; and an association of either of these markers with modulated SMC apoptosis.

We were unable to use the autopsy LAD samples to investigate these hypotheses because of extensive postmortem cell death and the decalcification of samples from individuals ≥45 years old. As an alternative, we obtained freshly processed carotid endarterectomies. Analysis of publicly deposited scRNA-seq data for carotid endarterectomies confirmed that the modulated SMC subtypes quantified in coronary atherosclerosis were also present in carotid plaques (**Suppl. Fig. 6**). Multiplex analysis for ACTA2/CD68/LUM and ACTA2/CD68/TNFRSF11B on carotid plaque sections (n=9) revealed that cells expressing only TNFRSF11B+ or only LUM+ accounted for 12% and 17% of all plaque cells, respectively (**Suppl. Fig. 7**), similar to the 12% and 13% in coronary fibroatheromas.

To investigate the co-localization of LUM+ cells with fibrotic connective tissue, we analyzed ACTA2/CD68/LUM-stained sections by fluorescence microscopy and subsequently stained the same sections with Aniline Blue (as a component of Masson’s trichrome) to identify collagen-rich tissue (**Fig. 5A**). Regions rich in fibrous tissue harbored many LUM+ cells, but LUM+ cells were also found outside these regions, and no significant differences were detected between LUM+ cell distribution within and outside fibrotic regions.

**Figure 5.**
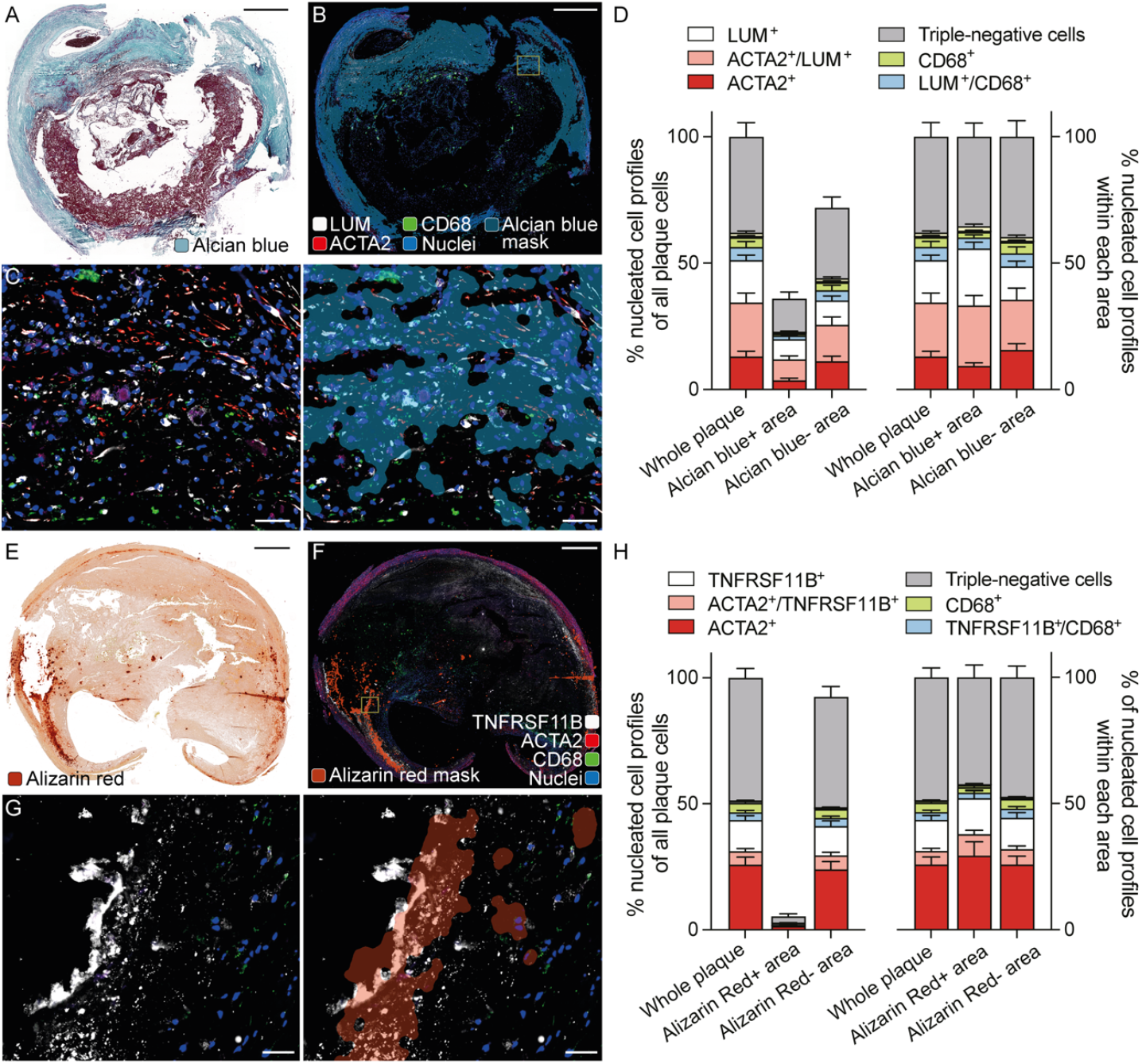
Association of modulated SMCs with fibrosis and calcification. **A,** Representative staining of carotid plaque with Aniline Blue (as part of Masson’s trichrome). **B-C,** ACTA2/CD68/LUM staining on the same section (before Masson’s trichrome), overlaid with a mask defined by high Aniline-blue signal in the Masson’s trichrome staining. Panel C shows the boxed region at higher magnification with and without the mask. **D**. Cellular composition inside and outside of the masked area, showing no differences according to fibrosity (n=9 carotid plaques analyzed). **E,** Representative staining with Alizarin Red. **F-G,** ACTA2/CD68/TNFRSF11B staining on an adjacent section, overlaid with a mask defined by Alizarin Red-detected calcium deposits. Panel G shows the boxed region at higher magnification with and without the mask. Note the positive staining of calcification granules. **H,** Cell phenotype (marker expression profile) inside and outside of the masked area, showing few cells in the calcified (Alizarin Red+) region and no differences in cell composition (n=9 carotid plaques analyzed). All bars show mean ± SEM. Scale bars 1000 μm (A,B,E,F) and 50 μm (C,G).

We used a similar approach to investigate the co-localization of TNFRSF11B+ cells with areas of calcification, this time staining with the ACTA2/CD68/TNFRSF11B antibody combination and Alizarin Red on adjacent sections (**Fig. 5B**). TNFRSF11B and Alizarin Red staining partly overlapped; however, few cells were found near calcifications, and calcified areas were not enriched in TNFRSF11B+ cells. Rather, the overlap in staining reflected the presence of TNFRSF11B on calcium deposits.

To determine the phenotypes of apoptotic cells in plaques, we performed triple staining with TUNEL, CD68, and either ACTA2, TNFRSF11B, or LUM, to distinguish between apoptotic cell remnants of macrophage, SMC, or modulated SMC origin (**Fig. 6**). Most TUNEL+ cells were negative for any marker, presumably due to marker-protein decay. Of the cell remnants for which an origin could be defined in TUNEL/CD68/ACTA2-stained sections, most were CD68+, whereas ACTA2+ apoptotic cells were uncommon. This is consistent with an earlier study that ascribed most cell death in plaques to macrophages based on this marker combination.^20^ However, antibody combinations including TNFRSF11B or LUM instead of ACTA2 identified contributions to apoptotic remnants by TNFRSF11B+ and LUM+ cells. This contribution was especially prominent in the case of LUM, with LUM+ cells contributing 35% and LUM+CD68+ cells 18% of all apoptotic remnants. Given that the scRNA-seq data identify LUM+CD68+ cells as a modulated SMC subtype, this finding indicates that modulated SMCs make a substantial and previously overlooked contribution to plaque apoptosis, accounting for as many apoptotic remnants as macrophages in advanced human atherosclerosis.

**Figure 6.**
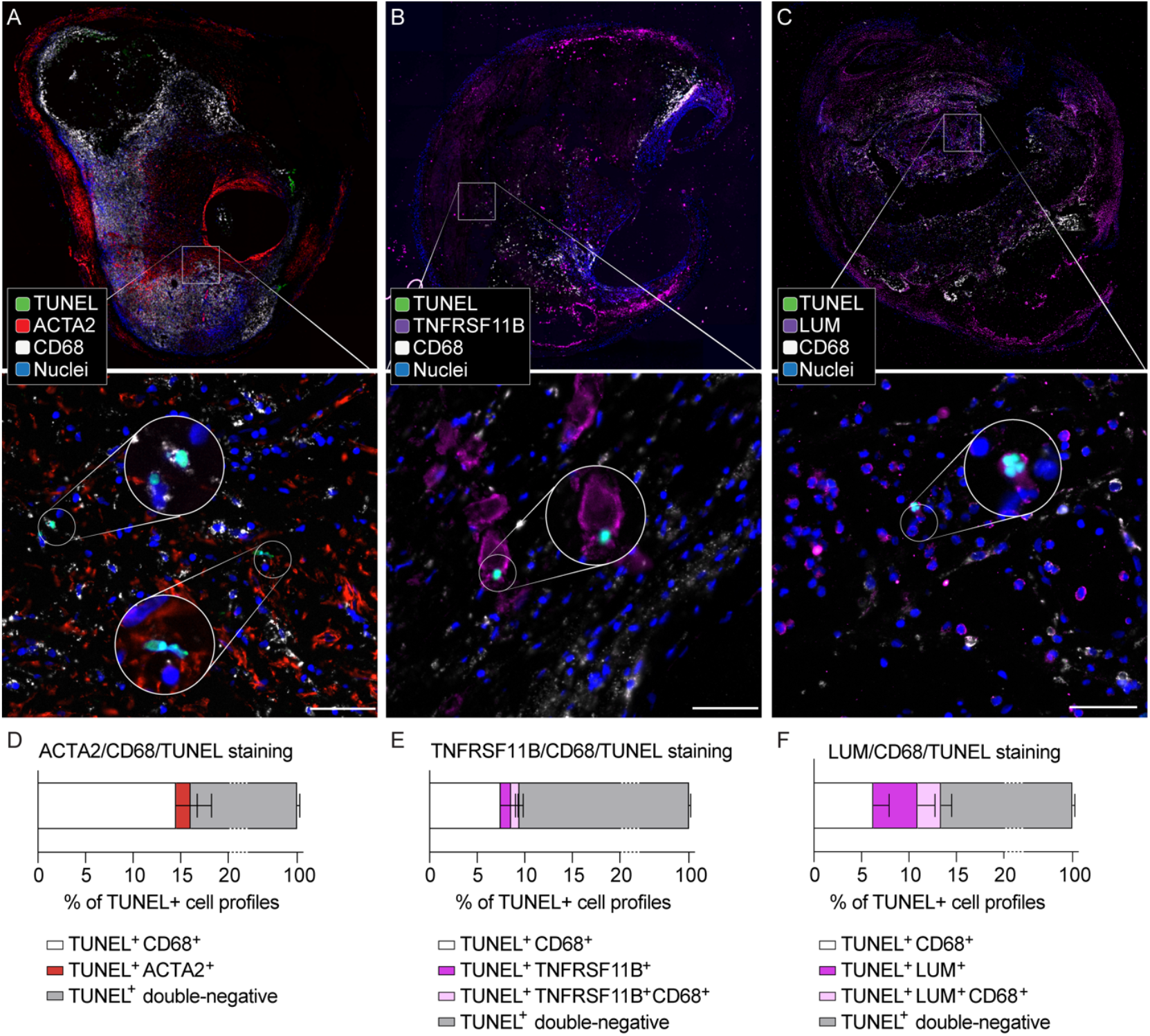
Contribution of modulated SMCs to plaque apoptotic cell remnants. **A-C.** Representative examples of TUNEL staining combined with immunofluorescence analysis for (A) CD68/ACTA2, (B) CD68/TNFRSF11B, and (C) CD68/LUM to determine the cellular origin of apoptotic cell remnants. Boxed regions are shown at higher magnification in the lower panels, and circled insets show cells at still higher magnification. **D-F**. Quantification of apoptotic cell phenotyping, showing that fully modulated LUM+ cells contribute substantially to apoptotic remnants for which an origin can be inferred (n=12 carotid plaques analyzed in each staining). Bars show mean ± SEM. Scale bars 50 μm.

## Discussion

The proliferation and modulation of local SMCs to alternative mesenchymal cell types is a key process in the growth of atherosclerotic lesions. The temporal pattern of SMC-derived cell recruitment has been studied in mouse models,^21,22^ but analysis of human atherosclerosis has been restricted to advanced plaques obtained from hearts explanted during transplantation procedures or from carotid endarterectomies.^1,2,6,23^ There has thus been a lack of knowledge about when during atherosclerosis the modulated cell types emerge. Furthermore, previous studies did not map the proximity of these cells to important plaque processes, such as fibrosis, calcification, and cell death, potentially useful information for defining their function in plaque development.

In the present study, we analyzed the temporal and spatial accumulation of modulated SMCs during human coronary atherosclerosis using two previously reported markers of modulated SMCs, TNFRSF11B and LUM,^1^ which were further characterized in the present study for their ability to identify specific modulated SMC populations. TNFRSF11B encodes the secreted glycoprotein osteoprotegerin, best known for its role in bone homeostasis.^19^ LUM encodes the extracellular matrix protein lumican, a small leucine-rich proteoglycan expressed by fibroblasts and previously implicated in myocardial fibrosis.^18^ Our re-analysis of the published scRNAseq data shows that LUM marks nearly all modulated SMCs that have lost ACTA2 expression, whereas TNFRSF11B marks an intermediate phenotype. Both markers showed high specificity for distinguishing SMC and modulated cells from other plaque cells, including macrophages. By establishing multiplex staining protocols and validated high-throughput image analysis techniques based on machine learning, we were able to phenotype the entire cell population in LAD and carotid plaques and link them to plaque stages and candidate plaque processes.

The multiplex immunofluorescence analysis showed that cells doubly positive for ACTA2 plus TNFRSF11B or for ACTA2 plus LUM are already present in normal arteries, whereas fully modulated TNFRSF11B+ and LUM+ cells lacking detectable ACTA2 expression are confined to atherosclerotic lesions and do not emerge until the fibroatheroma stage, coinciding with the appearance of the necrotic core. TNFRSF11B+ cells appeared to accumulate before LUM+ cells, consistent with a trajectory of modulation from contractile SMCs toward fibroblasts. That said, it remains unknown if SMC modulation is gradual or instead involves rapid changes in phenotype, which would amount to cells jumping from one site to another in UMAP plots.

Consistent with their presence in fibroatheromas, the appearance of LUM+ cells coincided with necrotic core formation, and these cells were enriched in the zone bordering the necrotic core in coronary atherosclerosis. Furthermore, LUM+ cells contributed a substantial proportion of apoptotic cell remnants in the carotid endarterectomy specimens. The contribution of SMCs to plaque cell death and necrosis has been a contested question. On the one hand, experimental studies have shown that forced apoptosis in plaque SMCs expands the necrotic core.^24^ On the other hand, human plaque studies phenotyping apoptotic cell remnants for ACTA2 and CD68, as in the present study, have generally identified macrophages as the dominant source.^20^ As our analysis indicates, this may be because traditional SMC markers such as ACTA2 do not detect the dying modulated SMC population. We found very few ACTA2+ apoptotic remnants, whereas modulated SMCs contributed on a par with macrophages, or even predominated if one considers LUM+ CD68+ cells as part of the SMC lineage. These findings are consistent with the transmission electron microscopy analysis of plaques by Kockx et al., which detected strangely shaped dying cells surrounded by basal laminae typical of SMCs.^25^ Interestingly, a recent large cellular GWAS study of apoptosis in cultured SMCs identified a locus that controls SMC apoptosis and is nominally associated with susceptibility to coronary artery disease.^26^ The abundance of LUM+ apoptotic cell remnants we detected near the necrotic core raises the possibility that the trackway from contractile to fully modulated SMCs may be associated with a progressive increase in the susceptibility to apoptosis.

Our examination of the association between modulated SMCs and other key pathogenetic mechanisms produced less conclusive results. Fibroblast-like LUM+ cells were present in fibrous plaque tissue but were not enriched at these locations compared with other regions. This might indicate that fibroblast-like cells with complete loss of ACTA2 expression are not required for the generation of fibrous tissue, which seems consistent with the wide expression of collagens by diverse mesenchymal cell types in human atherosclerotic plaque scRNA-seq data, including for ACTA2-expressing cells.^23,27^ Similarly, we found no evidence of physical proximity of TNFRSF11B+ cells to sites of calcification. However, secreted TNFRSF11B was localized in calcified regions, reminiscent of the detection of secreted TNFRSF11B on bone surfaces,^28^ where it reduces osteoclast activity by acting as a decoy receptor for the RANK ligand.^18^ In human and experimental atherosclerosis, calcification initiates in the necrotic core region,^29,30^ and unlike bone formation might not be tightly orchestrated by local cells. However, TNFRSF11B secreted by modulated SMCs may help to stabilize expanding areas of calcification. Alternatively, the detected extracellular TNFRSF11B could be derived from SMC-derived matrix vesicles, which have been implicated as nucleation sites of vascular calcification.^31,32^ The interpretation of previous studies in *Tnfrsf11b* knockout mice is complicated by excessive bone resorption, which may drive arterial calcification as a secondary effect.^33^ Further studies in mice with SMC-specific *Tnfrsf11b* deletion could be informative.

### Limitations

While the study focus was coronary atherosclerosis, we used freshly harvested carotid plaques for the apoptotic remnant analysis to avoid the distorting effect of artefactual postmortem apoptosis in the LAD autopsy material, which initial observations showed to be very extensive. Apoptosis is just one mode of cell death in atherosclerosis, which happens to be detectable because apoptotic remnants linger due to the suppression of efferocytosis.^34^ Other modes, such as necrosis, autophagic death, and pyroptosis, may also be important, but are difficult to detect because of their speed or lack of specific markers.^7^It should also be noted that the relative rate of apoptosis for different cell types can be under- or over-estimated if the remnants from different cell types are cleared with different efficiencies.^35^

## Conclusion

The present work shows that modulated SMCs, characterized by expression of TNFRSF11B or LUM without concomitant expression of ACTA2, accumulate only at the fibroatheroma stage of human coronary atherosclerosis. The fibroblast-like LUM-expressing cells, in particular, are enriched around the necrotic core and account for a substantial fraction of apoptotic remnants in human advanced plaques. Understanding the processes leading to full SMC modulation and cell death may thus present a useful strategy for developing plaque-stabilizing therapies that can limit necrotic core formation.

## Data Availability

All data produced in the present study are available upon reasonable request to the authors

## Acknowledgment

We thank the surgical team at the Department of Cardiothoracic and Vascular Surgery, Aarhus University Hospital, for carotid endarterectomy tissue collection; C. Torroja and F. Sanchez-Cabo of the CNIC Bioinformatics Unit for help with the initial analysis of public scRNA-seq data; members of the CNIC Histopathology Unit for excellent technical help; and S. Bartlett, CNIC, for English editing. Microscopy was partly conducted at the CNIC Microscopy & Dynamic Imaging Unit. The study was supported by grants from the Independent Research Fund Denmark (0134-00335B), the European Research Council through the European Union’s Horizon 2020 research and innovation program (grant no. 866240 to J.F.B.), the Novo Nordisk Foundation (NNF18OC0030688 to J.F.B.), the Spanish Ministerio de Ciencia, Innovación y Universidades with cofunding from the European Regional Development Fund (PID2019-108568RB-I00 to J.F.B. and IJC2020-044971-I to D.M.-C.), and Aarhus University Research Foundation (starting grant no. AUFF-E-201 9-723 to J.A.-J.). The CNIC is supported by the Instituto de Salud Carlos III, the Ministerio de Ciencia e Innovación, and the Pro-CNIC Foundation and is a Severo Ochoa Center of Excellence (grant no. CEX2020-001041-S, funded by MICIN/AEI/10.13039/501100011033).

