## Supplementary Figures and Tables for "Modulated smooth muscle cells accumulate late in human coronary atherosclerosis and are temporally and spatially linked to necrotic core formation"

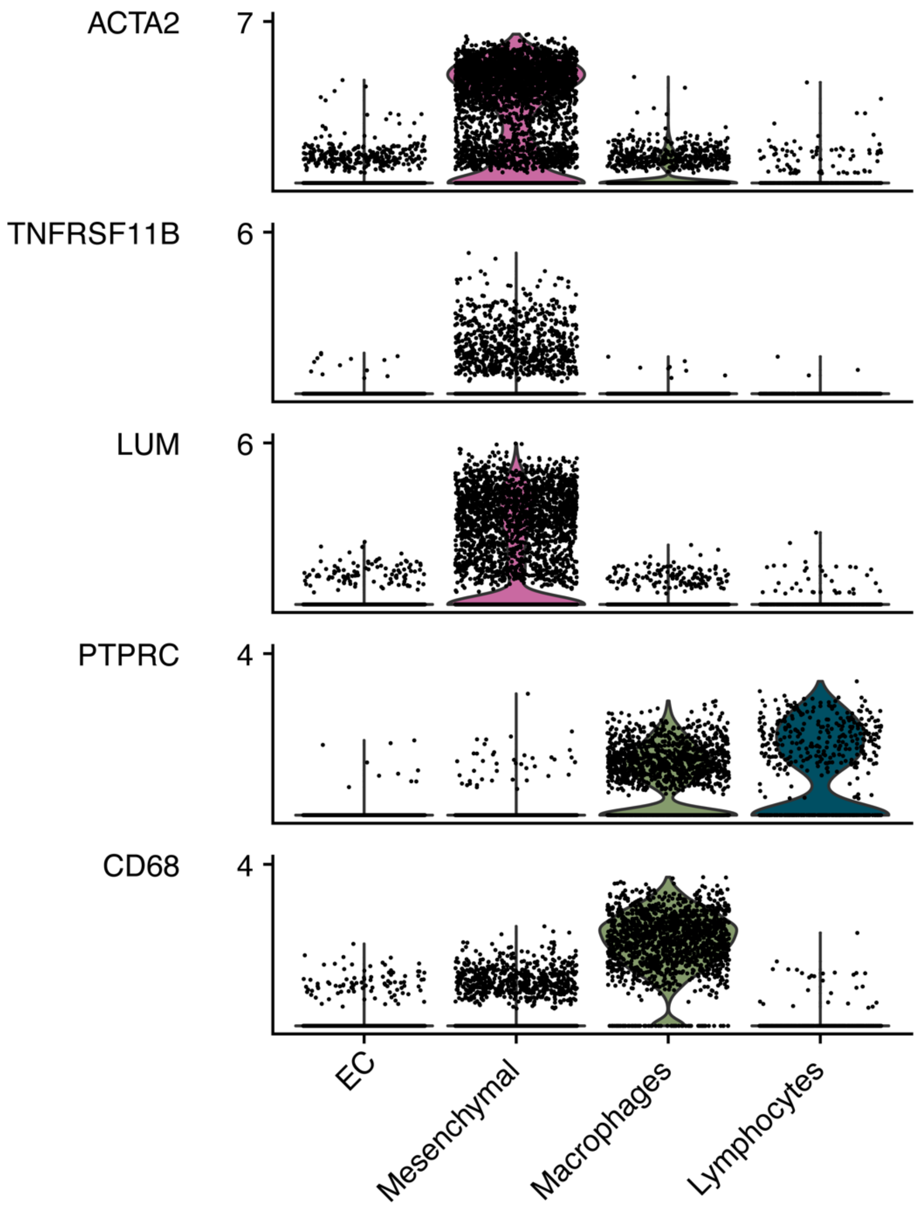

**Supplementary Figure 1. Specificity of cell type markers.** The violin plots show the normalized expression of the indicated marker genes in superclusters from coronary atherosclerosis scRNA-seq data (GSE131778).

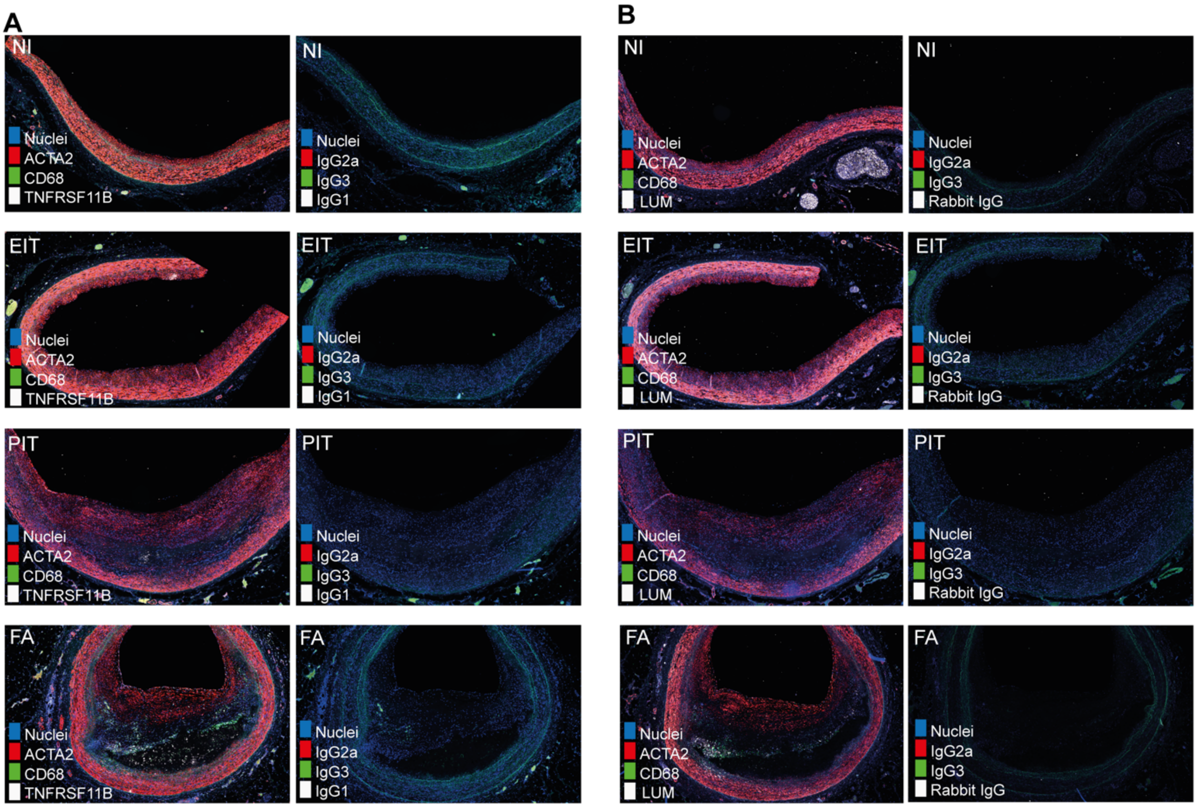

**Supplementary Figure 2. Isotype controls for Figure 3. A-B**. Panels show representative staining for (A) ACTA2/CD68/TNFRSF11B and (B) ACTA2/CD68/LUM on coronary artery sections with normal intima (NI), eccentric intimal thickening (EIT), pathological intimal thickening (PIT), and fibroatheromas (FA), accompanied by isotype-matched antibody staining on adjacent sections.

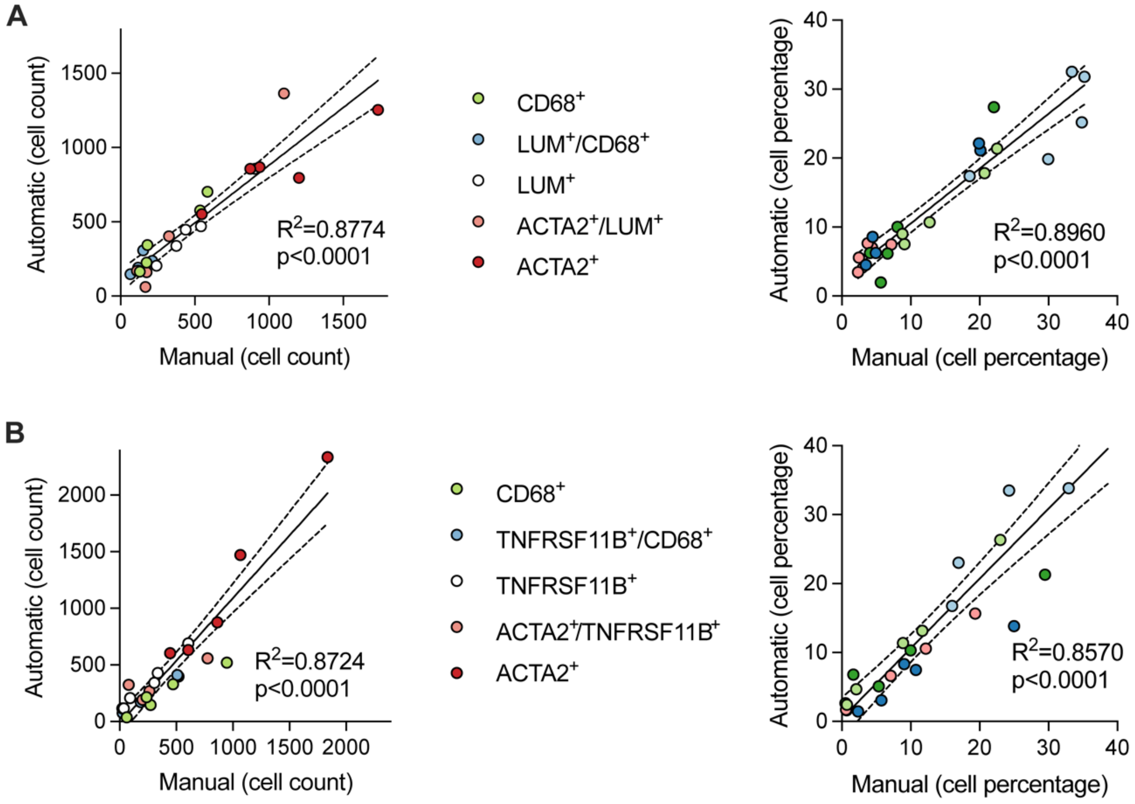

**Supplementary Figure 3. Validation of trained automatic cell phenotyping. A-B,** Comparison of automatic cell phenotyping in QuPath—based on the training of cell classifiers in specific plaque regions in a subset of sections—with manually curated cells, showing overall large agreement. Data represent cell counts (left) and percentages (right) in (A) 5 fibroatheroma sections stained for ACTA2/CD68/LUM and (B) 5 fibroatheroma sections stained for ACTA2/CD68/TNFRSF11B. $R^2$ correlation coefficients and p values were calculated by linear regression.

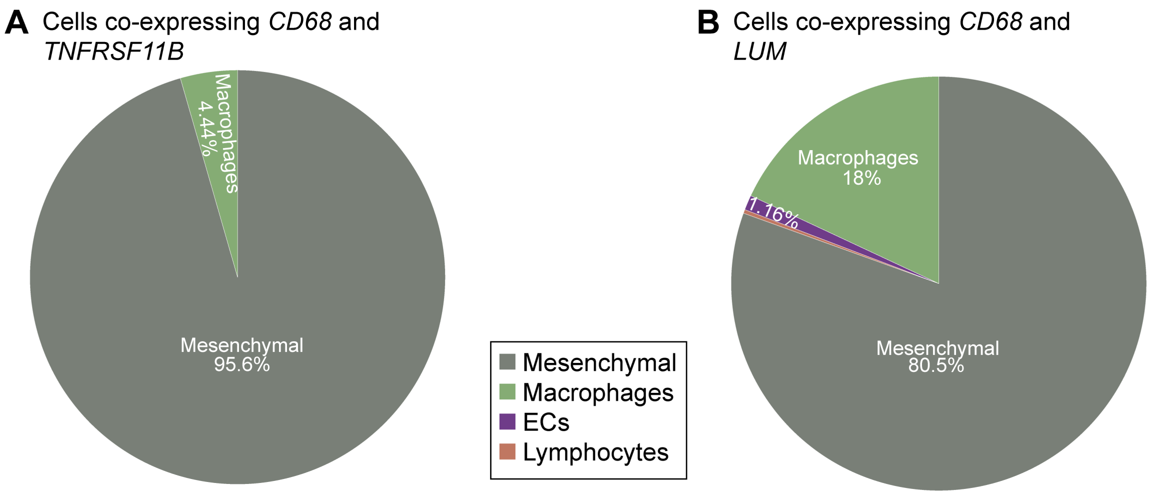

**Supplementary Figure 4. Identity of cells co-expressing modulated SMC and macrophage markers. A-B,** The likelihood of cellular identity was defined as the number of CD68/TNFRSF11B (A) or CD68/LUM (B) co-expressing cells in a cluster divided by the number of total co-expressing cells in coronary plaque scRNA-seq data (GSE131778).

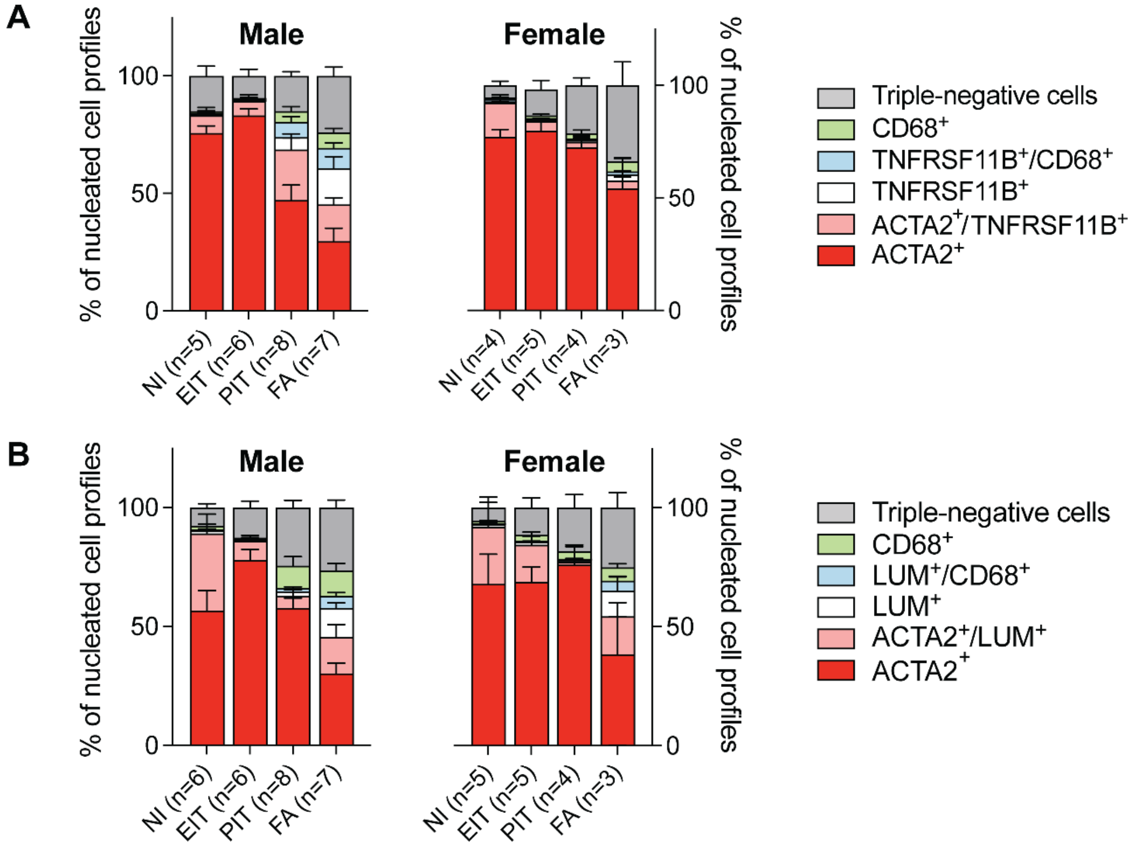

**Supplementary Figure 5. Cell composition in men and women during coronary atherogenesis. A-B.** Cell phenotype (marker expression profile) in sections stained for ACTA2/CD68/TNFRSF11B and for ACTA2/CD68/LUM in men and women. The plaque library included few advanced lesion stages from women, consistent with the slower atherosclerosis progression in women. Consequently, the pathological intimal thickening (PIT) and fibroatheroma (FA) categories contain fewer lesions from women. Nevertheless, the appearance of fully modulated LUM+ cells at the fibroatheroma stage is clear in both sexes. Bars show mean ± SEM. NI, normal intima. EIT, eccentric intimal thickening.

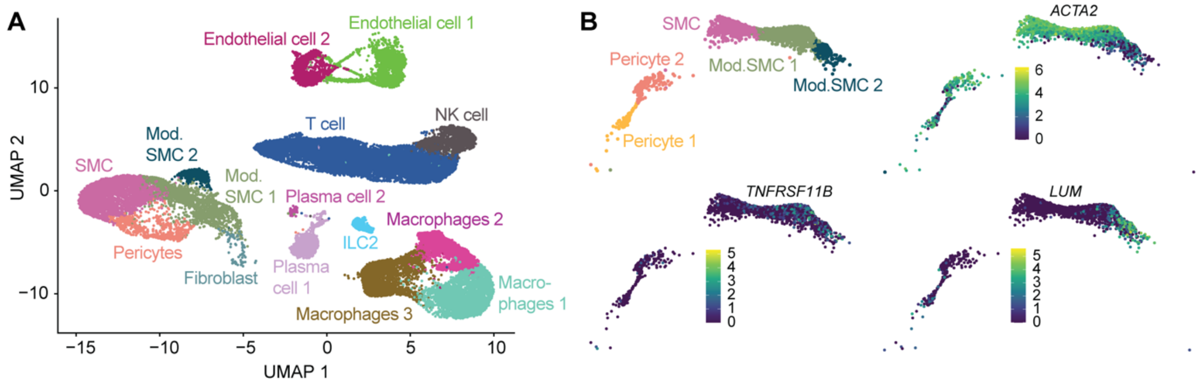

**Supplementary Figure 6. Markers of modulated SMCs in carotid plaque scRNA-seq data. A,** UMAP clustering of integrated public scRNA-seq data from carotid endarterectomies (GSE155512 and GSE159677). **B,** Projection of carotid cells from the mesenchymal supercluster to the UMAP for the coronary data (GSE131778; see Figure 2). Expression of *ACTA2*, *TNFRSF11B*, and *LUM* places cells on an axis of phenotypic diversity from contractile ACTA2+ SMCs to fibroblast-like LUM+ cells, similar to the pattern detected in the coronary atherosclerosis scRNA-seq data.

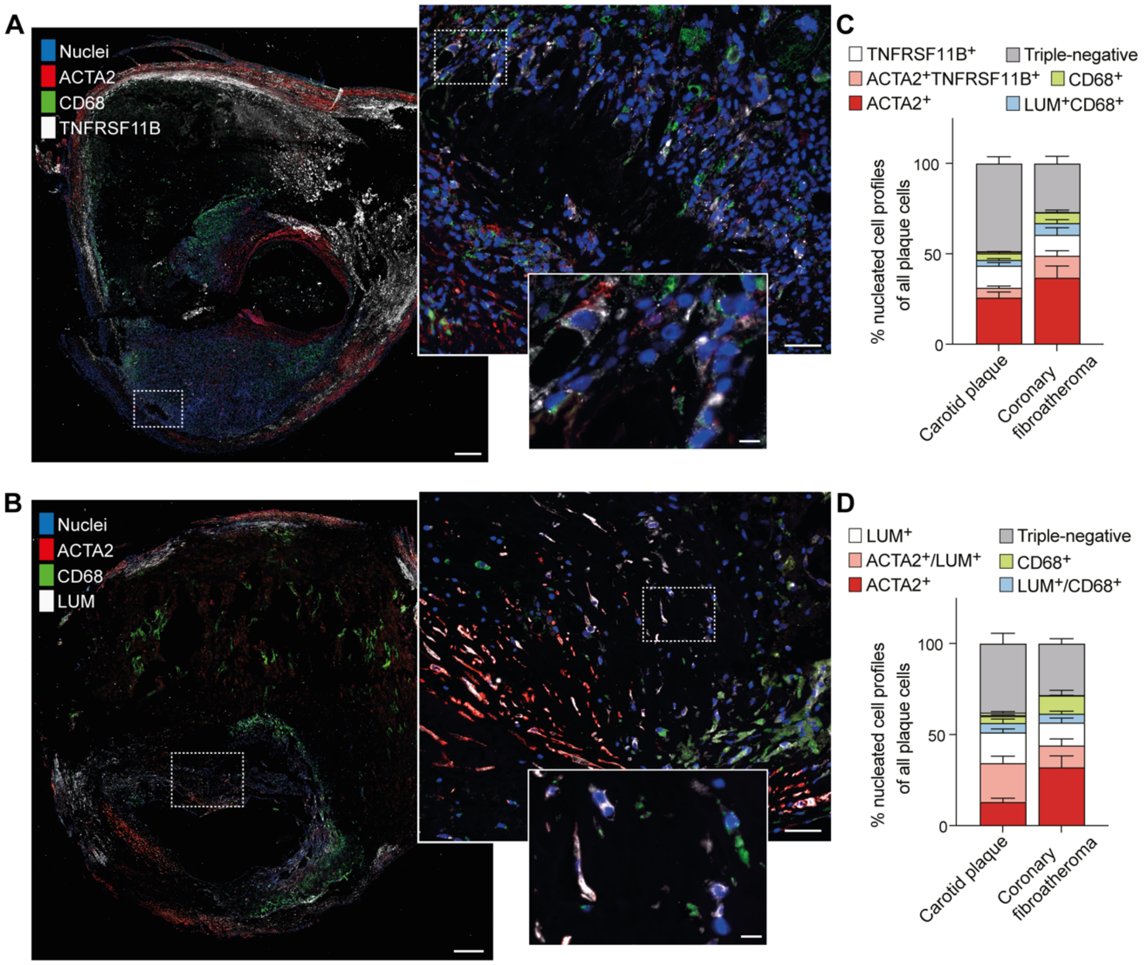

**Supplementary Figure 7. Modulated SMCs in carotid plaques. A-B**, Representative examples of carotid plaque sections stained for ACTA2/CD68/TNFRSF11B or ACTA2/CD68/LUM. The boxed areas are shown at higher magnification. Scale bars, 500 µm, 50 µm, and 10 µm in the low, intermediate, and high-magnification images, respectively. **C-D**, Cell phenotype (marker expression profile) in carotid plaques shown alongside that in coronary fibroatheromas for comparison (n=9 carotid plaques analyzed in each staining). Data are shown as mean ± SEM is shown.

**Supplementary Table 1. Primary antibodies used in the study.**

| Target | Clone/ catalogue # | Host | Source | Working dilution |
|---|---|---|---|---|
| ACTA2 | 1A4/ M0851 | Mouse $IgG_{2a}$ | Dako | 1:200 |
| CD45 | 2B11 + PD7/26 / M0701 | Mouse $IgG_1$ | Dako | 1:50 |
| CD68 | PG-M1/ M0876 | Mouse $IgG_3$ | Dako | 1:50 |
| LUM | EPR8898(2)/ ab198974 | Rabbit monoclonal | Abcam | 1:50 |
| OPG | [98A1071]/ NB-100-56505 | Mouse $IgG_1$ | Novus Biologicals | 1:100 |
| $IgG_{2a}$ | CLCMG2A00 | Mouse | Cerderlane lab | conc. matching primary |
| $IgG_1$ | [B11/6] / ab91353 | Mouse | Abcam | conc. matching primary |
| $IgG_3$ | 14474282 | Mouse | ThermoFisher | conc. matching primary |
| IgG | [DA1E]/ 3900S | Rabbit | Cell Signaling Technology | conc. matching primary |

**Supplementary Table 2. Secondary antibodies used in the study**

| Target | Clone/ catalogue # | Conjugate | Host | Source | Working dilution |
|---|---|---|---|---|---|
| Mouse $IgG_3$ | A21151 | Alexa Fluor® 488 | Goat | ThermoFisher | 1:500 |
| Mouse $IgG_3$ | 115-605-209 | Alexa Fluor® 647 | Goat | Jackson ImmunoResearch | 1:500 |
| Mouse $IgG_{2a}$ | A21134 | Alexa Fluor® 568 | Goat | ThermoFisher | 1:500 |
| Mouse $IgG_1$ | A21240 | Alexa Fluor® 647 | Goat | ThermoFisher | 1:500 |
| Mouse $IgG_1$ | A21124 | Alexa Fluor® 568 | Goat | ThermoFisher | 1:500 |
| Rabbit IgG (H+L) | A11011 | Alexa Fluor® 568 | Goat | ThermoFisher | 1:500 |
| Rabbit IgG (H+L) | A21245 | Alexa Fluor® 647 | Goat | ThermoFisher | 1:500 |
